# A randomised controlled trial of the effect of intra-articular lidocaine on pain scores in inflammatory arthritis

**DOI:** 10.1101/2023.10.30.23297628

**Authors:** Z. Rutter-Locher, S. Norton, F. Denk, S. McMahon, L.S. Taams, K. Bannister, B. Kirkham

## Abstract

**Background:** Chronic pain in inflammatory arthritis (IA) reflects a complex interplay between active disease in a peripheral joint and central pro-nociceptive mechanisms. Since intra-articular lidocaine may be used to abolish joint-specific peripheral input to the central nervous system, we aimed to validate its use as a clinical tool to identify those IA patients whose pain likely incorporates centrally mediated mechanisms.

**Methods:** In this two-armed randomised placebo-controlled trial, IA patients requiring an intra-articular steroid injection were 1:1 randomised to additionally receive intra-articular lidocaine or control (0.9% saline). Pain numerical rating scale (NRS) scores were collected at baseline and 3, 5, and 10 minutes post injection. Between group differences in NRS scores at each post-randomisation assessment were estimated using linear mixed-models. Heterogeneity in lidocaine effect was evaluated by baseline painDETECT (grouped ‘high’ (>18) or ‘low’ (≤18)). Analysis in a second cohort validated the painDETECT analysis and included additional markers of centrally mediated pain.

**Results:** The placebo effect of intra-articular injection was low. Post lidocaine injection, those in the high painDETECT group had an NRS score 2.2 points higher than those in the low painDETECT group (p=0.03). In the replication sample, post lidocaine NRS scores were significantly higher in those with a high painDETECT score, fibromyalgia, and low-pressure pain threshold at the trapezius (p=0.002, p=0.001, p=0.005 respectively).

**Conclusion:** Persistent high pain post intra-articular lidocaine injection could potentially be used as an indicator of pro-nociceptive mechanisms that are centrally mediated, informing centrally-targeted analgesic strategies.

## Introduction

Chronic pain in inflammatory arthritis (IA) arises because of a complex interplay between active disease in peripheral joints and central pain processing mechanisms^1^. This interplay differs between patients, meaning that identifying the predominant underlying pain-driving mechanism, while difficult, is crucial if optimal pharmacotherapies are to be prescribed. Currently, peripherally mediated pain in IA is inferred indirectly by examining the degree of inflammation or joint damage (as assessed by ultrasound). In contrast, although validated questionnaires and quantitative sensory testing (QST) can be used to determine the trait versus state nature of the pain experience and the integrity of pain processing circuits respectively, there is no ‘gold standard’ method to infer the presence of centrally mediated pain in IA despite its predicted prevalence in up to 40% of IA patients^2^. Pinpointing peripheral versus central nervous system contributions to the pain state could aid phenotyping of pain, where stratifying patients into appropriate mechanism-based pain ‘cohorts’ is a key goal and could aid targeted analgesic treatment for the individual^3,4^.

Lidocaine blocks voltage-gated sodium channels leading to a reversible blockade of action potential propagation in peripheral nerves at the site of injection. Therefore, administration of intra-articular lidocaine in a diseased joint should largely abolish the peripheral drive from that joint and, theoretically, lead us towards the possibility of uncoupling centrally versus peripherally mediated pain. Lidocaine administration was previously validated to identify peripherally mediated pain in syndromes including peripheral neuropathic pain, phantom limb pain and bladder pain syndrome^5–7^. We previously demonstrated that patients with bladder pain syndrome could be categorised as having predominantly peripherally versus centrally mediated pain following intravesical lidocaine infiltration^7^. Interestingly, those patients who did not respond to lidocaine (i.e., pain scores were not reduced by >50%) were 2.7 times more likely to experience other central sensitivity syndromes, indicative of centrally mediated pain^7^.

In the present study, we hypothesised that an IA patient population could be stratified into two cohorts based on their response to intra-articular lidocaine. We hypothesised that pain numerical rating scale (NRS) scores would reduce in all patients due to lidocaine blocking a large component of the peripherally mediated pain, but that those with contributing centrally mediated pain would report higher ongoing post injection pain. By comparing the response in pain NRS to intra-articular injection of lidocaine versus a control injection of saline, our primary objective was to identify a change in NRS potentially caused by a placebo effect. Our secondary objective was to group patient responses to lidocaine injection according to their painDETECT scores (a validated questionnaire used to assess the presence of possible neuropathic like pain, as a proxy marker of centrally mediated pain^8^), hypothesising that those patients in the high painDETECT (score >18) group would, post-lidocaine injection, rate their pain higher than those patients in the low painDETECT (score ≤18) group. Finally, we replicated and extended the analysis of our secondary objective in a second patient cohort using multiple markers of centrally mediated pain including fulfillment of fibromyalgia criteria and dynamic quantitative sensory testing outcomes.

## Methods

### Trial design

This two-armed parallel group randomised controlled trial was approved by Yorkshire & The Humber-Sheffield research ethics committee (REC reference 22/YH/0051). All patients gave written informed consent. This study was pre-registered on www.clinicaltrials.gov prior to first patient enrollment (Identifier NCT05302232, Unique protocol ID 311106) and has been designed and reported in line with the CONSORT 2010 guidelines and checklist ^9,10^.

### Participants

Patients with a diagnosis of IA, including but not limited to rheumatoid arthritis and psoriatic arthritis, with a numerical rating scale (NRS) pain score > 3/10 and who required an intra-articular steroid injection as recommended by the direct care team were recruited from Guy’s Hospital Rheumatology department. We excluded those with underlying joint damage identified on routine x-ray, those under 18 years of age and those requiring shoulder or proximal interphalangeal joint injection.

### Randomization and Interventions

Patients were block randomized in a 1:1 allocation ratio, using the online randomization service sealed envelope (Sealedenvelope.com), to receive either intra-articular 1% lidocaine plus steroid or, as a placebo control, 0.9% saline plus steroid. Patients only were blinded to study group. For pragmatic reasons, the outcome assessor also administered the injections and was thus unblinded. Standardised amounts of steroid and lidocaine were administered for each joint (Supplementary Table 1).

### Outcomes

Demographics including age, gender and diagnosis were collected. Pain scores (NRS 0-10) at rest in the chosen joint were collected prior to injection and at 3, 5 and 10 minutes post injection, when the lidocaine would be expected to have an analgesic effect ^11^. Since the steroid is slower acting than lidocaine, it should not have a beneficial effect on pain scores within ten minutes. Improvement in pain rating within 10 minutes by patients who received steroid only would therefore be due to placebo effect and random variation alone. Needle placement within the joint was confirmed by fluid aspiration.

Participants completed the painDETECT questionnaire at baseline. This assesses possible neuropathic like pain as a proxy for centrally mediated pain and has sensitivity and specificity of 84%, using clinician-assessed diagnosis of neuropathic pain as the gold standard^12^. For the purposes of this study painDETECT scores were grouped into high (>18) or low (≤18) likelihood of neuropathic like pain.

### Sample size

Using our pilot data, we estimated that 80 patients would be required (40 in each group) to achieve 80% power to detect a difference in NRS of at least 1.5 points between the experimental group (lidocaine) compared to the control group at the 5% alpha level. This was based on the expectation that we would perform an ANCOVA and on assuming a SD of 2.9 for the NRS, and a pre-post NRS correlation of r= 0.4. A planned interim analysis was conducted at a total sample size of 50 (i.e. 25 per group). To account for this analysis the critical alpha level was set at p=0.03 based on the Pocock method for alpha control (i.e. multiple testing) in sequential analyses. The interim analysis met the stopping criteria and thus recruitment was halted early and the final sample size was 51.

### Statistical methods

Means with standard deviation (SD) are used to describe continuous demographics and painDETECT scores. For the primary objective, a linear mixed effects model was used to estimate between group differences (lidocaine versus control) at each of the post-intervention time points, adjusted for the baseline level of the outcome. Specifically, group allocation and assessment time were included as categorical variables using dummy coding along with a time-by-group interaction terms. Baseline NRS was included as a covariate. A random intercept was included to account for the repeated assessments within individuals. Further analysis extended the model by including a dummy coded covariate for high versus low painDETECT score and three-way interaction terms between painDETECT with group and time. This allows for the interrogation of lidocaine effect heterogeneity by estimating specific lidocaine treatment effects for those with high versus low painDETECT scores at each time point. Analysis was performed in Stata V 17.1. For the between group difference (lidocaine versus control), the significance threshold was set at p<0.03 due to the sequential design used and p<0.05 for all other analyses.

### Extended analysis in second patient cohort

Supplementary analysis was performed using data from 40 patients enrolled in the “Pain phenotypes and their Underlying Mechanisms in Inflammatory Arthritis” study (PUMIA), which received approval from Bromley research ethics committee and the Health Research Authority (REC 21/LO/0712). All patients gave written informed consent.

Inclusion and exclusion criteria mirror those in the main RCT and the same visit schedule and outcomes were collected. All patients received intra-articular 1% lidocaine along with steroid injection. In addition to collection of the painDETECT questionnaire, fibromyalgia status was determined according to the 2016 revision of the ACR 2010 modified fibromyalgia diagnostic criteria^13^ and quantitative sensory testing (QST) was performed. QST included evaluation of pressure pain thresholds at a non-articular site as a marker of widespread pain sensitisation (the bilateral trapezius, taken as an average), temporal summation of pain (TSP) and conditioned pain modulation (CPM) using cuff algometry, as detailed in previous publications ^14^

Given the nature of the supplementary analysis, no a-priori power calculation was performed. The sample size provides 73% power to detect a correlation of at least 0.4 based at the 5% (two-sided) significance level.

A linear mixed effects model estimated between group differences in NRS by painDETECT group (high vs low) at 3, 5 and 10 minutes post lidocaine injection. Specifically, painDETECT group and time were included as dummy coded variables along with interaction terms. The analyses were repeated replacing painDETECT group with other related pain variables: positive or negative for fulfillment of fibromyalgia criteria, pressure pain threshold (PPT) high vs low group (above or below median PPT at the trapezius), positive or negative for facilitated TSP (using ratio >2.48 to represent facilitated TSP^14^) and responder or non-responder to CPM (using CPM response >20% of the baseline pressure tolerance threshold to represent responders and CPM response <20% of the baseline pressure tolerance threshold to represent non-responders^14^). Using these additional groupings, positive fulfillment of fibromyalgia criteria, PPT low group, positive for facilitated TSP and non-responder to CPM were deemed to be proxy measures of centrally mediated pain.

## Results

### Recruitment and participant flow

Fifty-one patients were recruited between April and October 2022, 26 in the treatment (lidocaine) group and 25 in the control group. No participants were lost or excluded post randomisation as randomisation and intra-articular injection occurred on the same study visit.

### Baseline data

Demographics are shown in Table 1. 64.7% were female and mean age was 53.4 years with a large range of 26-84 years, in keeping with the disease demographics. Most participants had rheumatoid arthritis (61%) or psoriatic arthritis (19.6%). The most common joints injected were the knee (57%) or wrist (28%). Pre-injection NRS were high, as would be expected for patients requiring joint injection.

**Table 1:**
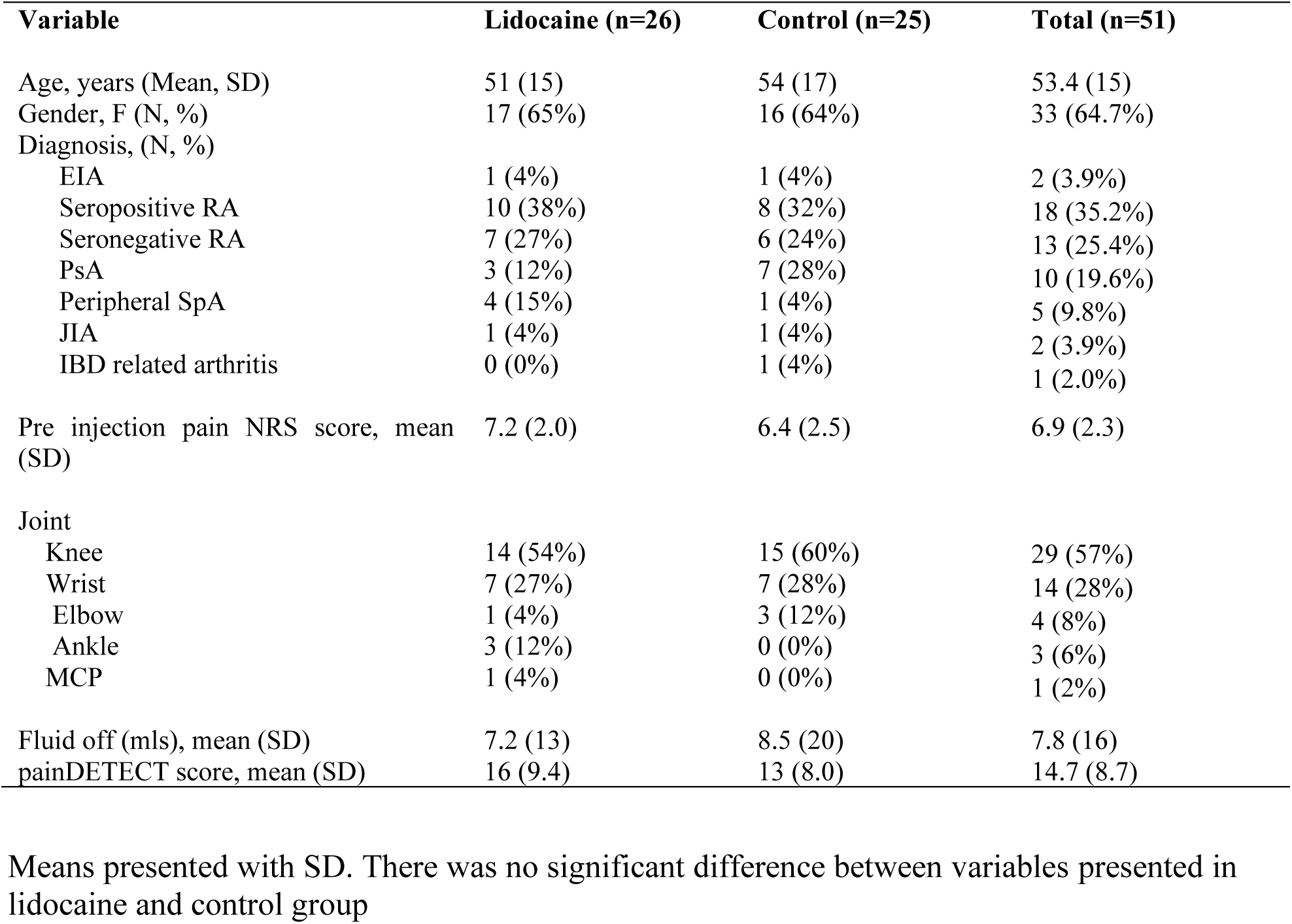
Demographics of study population. Means presented with SD. There was no significant difference between variables presented in lidocaine and control group.

| Variable | Lidocaine (n=26) | Control (n=25) | Total (n=51) |
| --- | --- | --- | --- |
| Age, years (Mean, SD) | 51 (15) | 54 (17) | 53.4 (15) |
| Gender, F (N, %) | 17 (65%) | 16 (64%) | 33 (64.7%) |
| Diagnosis, (N, %) |  |  |  |
| EIA | 1 (4%) | 1 (4%) | 2 (3.9%) |
| Seropositive RA | 10 (38%) | 8 (32%) | 18 (35.2%) |
| Seronegative RA | 7 (27%) | 6 (24%) | 13 (25.4%) |
| PsA | 3 (12%) | 7 (28%) | 10 (19.6%) |
| Peripheral SpA | 4 (15%) | 1 (4%) | 5 (9.8%) |
| JIA | 1 (4%) | 1 (4%) | 2 (3.9%) |
| IBD related arthritis | 0 (0%) | 1 (4%) | 1 (2.0%) |
| Pre injection pain NRS score, mean (SD) | 7.2 (2.0) | 6.4 (2.5) | 6.9 (2.3) |
| Joint |  |  |  |
| Knee | 14 (54%) | 15 (60%) | 29 (57%) |
| Wrist | 7 (27%) | 7 (28%) | 14 (28%) |
| Elbow | 1 (4%) | 3 (12%) | 4 (8%) |
| Ankle | 3 (12%) | 0 (0%) | 3 (6%) |
| MCP | 1 (4%) | 0 (0%) | 1 (2%) |
| Fluid off (mls), mean (SD) | 7.2 (13) | 8.5 (20) | 7.8 (16) |
| painDETECT score, mean (SD) | 16 (9.4) | 13 (8.0) | 14.7 (8.7) |

### Placebo effect of intra-articular injection is low

The mean pain NRS score was 3.5 points lower than baseline at 5 minutes post injection in the intra-articular lidocaine group and 1.2 points lower than baseline at 5 minutes post injection in the steroid only placebo control group (Figure 1A). Decomposition of the total change from baseline indicated that a 2.8 point difference (81% of the total effect) was due to the treatment effect of lidocaine and the remaining 0.7 points (19%) placebo effect.

**Figure 1.**
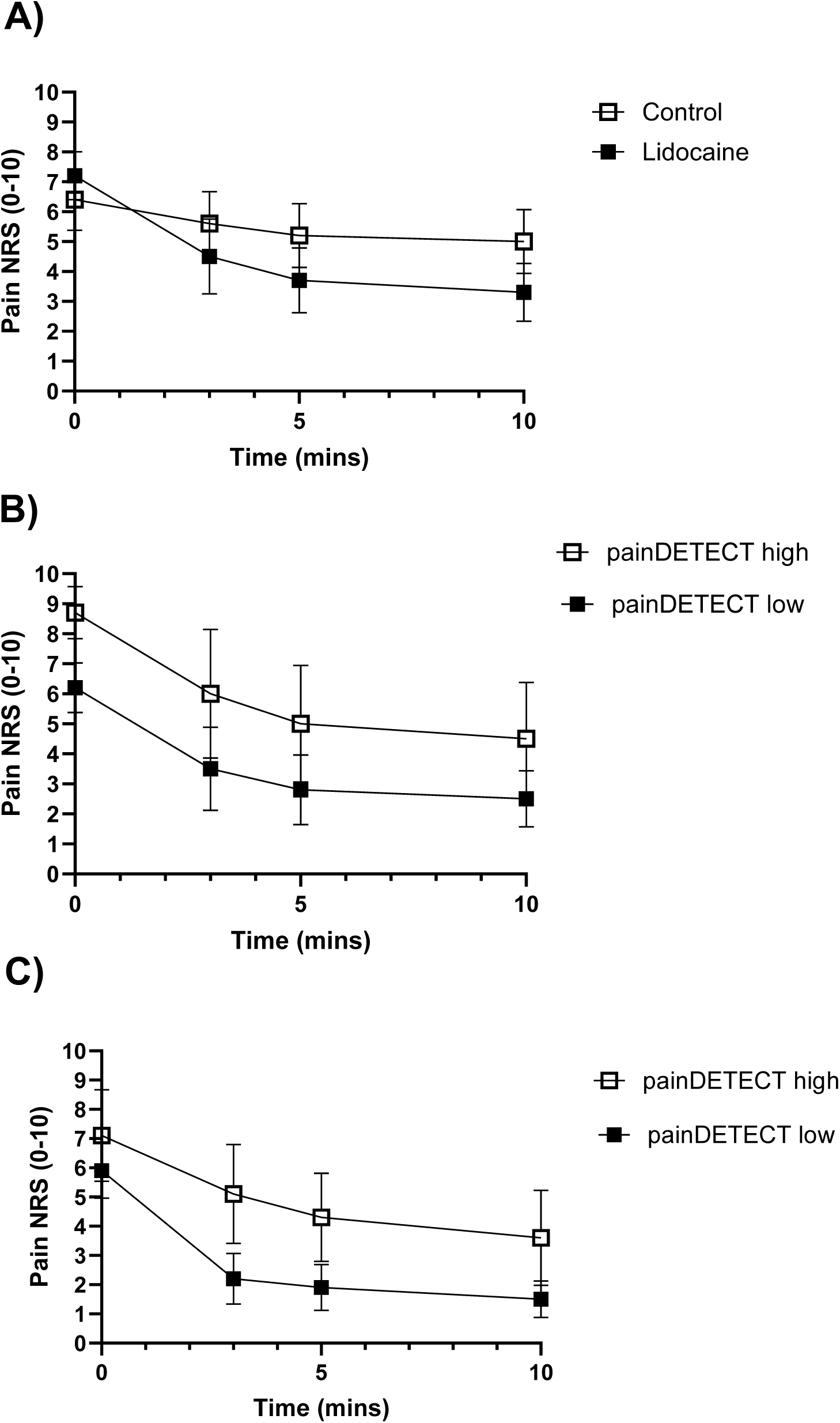
NRS response at 3,5 and 10 minutes post to intra-articular injection (A) Pain NRS score pre and post lidocaine versus steroid only control injection in RCT group (mean ± 95% CI). (B) Pain NRS score pre and post lidocaine, grouped by painDETECT low/high in first patient cohort (RCT group) (mean ± 95% CI). (C) Pain NRS score pre and post lidocaine, grouped by painDETECT low/high in second validation cohort (PUMIA group).

Linear mixed effect regression, adjusting for baseline pain NRS scores, indicated that the lidocaine group had significantly lower NRS scores on average across the post-injection time points, compared to the control group (main-effect p=0.002). The adjusted mean differences were observed to be significant at each of the three assessment points with an increasing effect over time (Table 2).

**Table 2:** Adjusted Mean Differences for Lidocaine Compared to Control group at Different Time Points (Raw and Standardized Values)

| Time (mins) | Raw Marginal Effect | Standardized Marginal Effect | Significance |
| --- | --- | --- | --- |
| 3 | 1.79 | 0.65 | p = 0.002 |
| 5 | 2.14 | 0.78 | p < 0.001 |
| 10 | 2.40 | 0.87 | p < 0.001 |

### Patients with a high painDETECT score report higher ongoing pain following intra-articular lidocaine

Regardless of high (>18) or low (≤18) pre-injection painDETECT scores, a comparable reduction in pain NRS scores were reported by patients post lidocaine injection (Figure 1B, 4.2 versus 3.7 points respectively), and post placebo injection (Supplementary Figure 1, 1.1 versus 1.4 points respectively). This potentially indicates a similar level of peripherally driven nociceptive pain (presumably generated by joint inflammation) in all patients. The mean NRS score 5 minutes post intra-articular lidocaine was 5 in the high painDETECT group and 2.8 in the low painDETECT group. On average, across all three post-injection assessments, those in the high painDETECT group had a NRS score that was 2.2 points higher than those in the low painDETECT group (p=0.025, Table 3 and Supplementary Table 3). A sensitivity analysis was run controlling for baseline NRS scores; this reduced the average difference in NRS between the groups across all three post-injection assessments to non-significant (0.5, p=0.49).

**Table 3:**
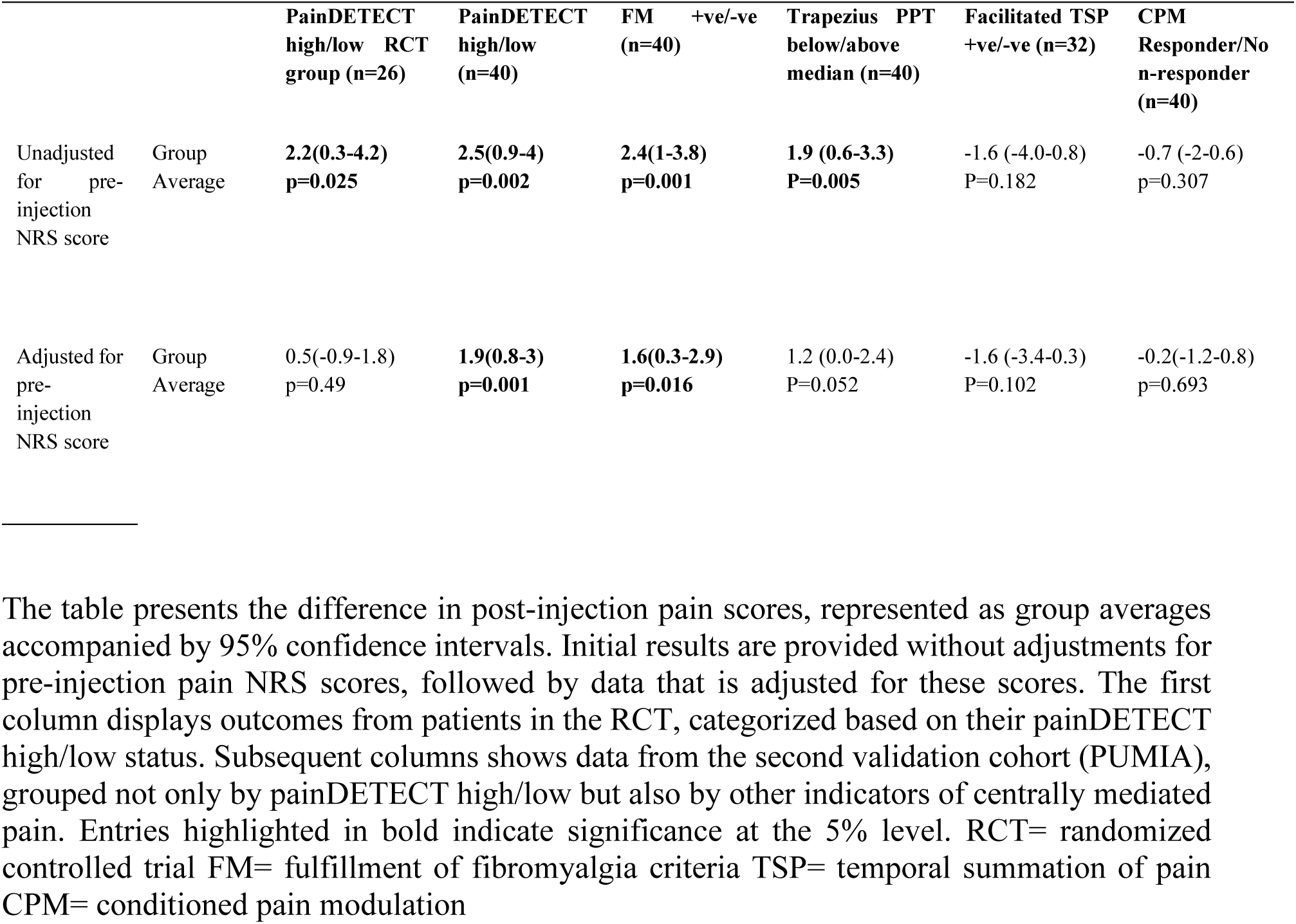
Results of the linear mixed effect model. The table presents the difference in post-injection pain scores, represented as group averages accompanied by 95% confidence intervals. Initial results are provided without adjustments for pre-injection pain NRS scores, followed by data that is adjusted for these scores. The first column displays outcomes from patients in the RCT, categorized based on their painDETECT high/low status. Subsequent columns shows data from the second validation cohort (PUMIA), grouped not only by painDETECT high/low but also by other indicators of centrally mediated pain. Entries highlighted in bold indicate significance at the 5% level. RCT= randomized controlled trial FM= fulfillment of fibromyalgia criteria TSP= temporal summation of pain CPM= conditioned pain modulation

### Patients with a high painDETECT score, fibromyalgia and low non articular PPT report higher ongoing pain following intra-articular lidocaine, in a second patient cohort

Forty patients were included in the replication (validation) analysis using the PUMIA cohort. Demographics are shown in Supplementary Table 4. 73% were female and mean age was 52.7 years. Most participants had rheumatoid arthritis (83%). The most common joints injected were the wrist (58%) or knee (23%). Median PPT at the trapezius was 2.1 Kg/cm^2^.

Regardless of high (>18) or low (≤18) pre-injection painDETECT scores, a comparable reduction in NRS scores was again reported by patients post lidocaine injection (Figure 1C). The mean NRS score at 5 minutes post intra-articular lidocaine was 4.3 in the high painDETECT group and 1.9 in the low painDETECT group. Based on a mixed effects model, those in the high painDETECT group had a post lidocaine injection NRS score that was 2.5 points higher than those in the low painDETECT group (Table 3, main effect p=0.002) and NRS scores were significantly higher in the high painDETECT group at each post intervention time point (Supplementary Table 3). Sensitivity analysis, controlling for pre-injection NRS scores, revealed that, although less attenuated, in this second cohort there was still a significant difference between pain scores post intra-articular lidocaine injection in those with high versus low painDETECT (Table 3).

Further analysis considered other markers of central pain processing. Average post injection NRS scores were significantly higher in those fulfilling fibromyalgia criteria, compared to those who did not, both when adjusting or not adjusting for pre-injection pain scores (Figure 2A and Table 3). Average post injection NRS scores were also significantly higher in those with low, versus high, PPT at the trapezius, when not adjusting for pre-injection NRS scores, but this effect did not survive correction for pre-injection NRS scores (Figure 2B and Table 3). Post injection NRS scores were also numerically higher in those with facilitated TSP, compared with not facilitated TSP, and those who were non-responders to CPM compared to responders but this was not significant (p= 0.182 and p=0.307 respectively, Figure 2C and 2D and Table 3). Ratings at 5 minutes post intra-articular lidocaine in those who had a high a painDETECT and who fulfilled fibromyalgia criteria consistently exceeded 4/10 (Supplementary Table 2).

**Figure 2.**
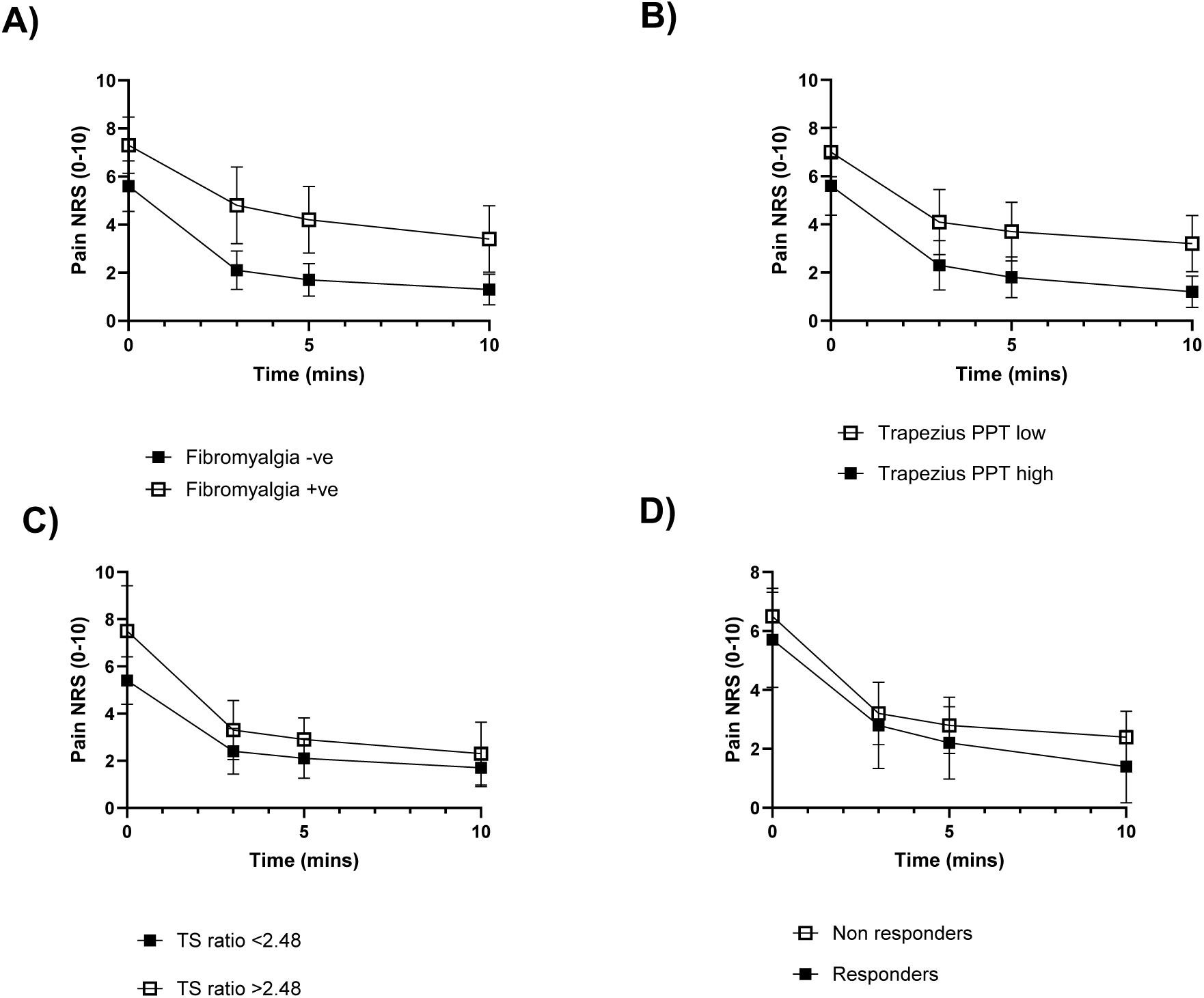
Drop in pain NRS score pre and post lidocaine in PUMIA patients, grouped by A) Fulfilment of fibromyalgia criteria +ve/-ve, (p=0.001) B) pressure pain threshold (PPT) high vs low group (above or below median PPT at the trapezius) (p=0.005), C) Fulfilment of temporal summation of pain (TS) ratio >2.48 +ve/-ve,(p=0.182) or D) Responder or Non-responder to conditioned pain modulation (CPM) pressure tolerance threshold (p=0.307) (Mean ± 95%CI).

## Discussion

We found that in patients with IA, the placebo response to an intra-articular injection was low, suggesting that the beneficial effect of intra-articular lidocaine on pain scores (as measured using the NRS) was mainly the result of a peripheral neurological action, rather than a distinct placebo response. The placebo response, widely studied in pain science ^15,16^, is influenced by internal factors such as patient expectation, emotions, and past experiences, as well as external factors such as verbal suggestions, social cues, and body language ^17^. It has a well-defined biological foundation that includes not only the autonomic and neuroendocrine systems but also modulatory processes involving the prefrontal cortex and the axis of the periaqueductal grey (PAG), rostroventral medulla (RVM), and spinal cord ^18^. Given this complexity, it was vital to evaluate whether any element of the pain-reducing effect of an intra-articular injection was governed by a placebo mechanism.

We observed a reduction in pain NRS score following lidocaine injection, indicating a significant component of pain that was peripherally mediated. Lidocaine, as a non-selective voltage gated sodium channel blocker, prevents depolarization and action potential propagation in all peripheral nerve fibres (including motor, sensory and autonomic) at the site of injection^7^. When administered into a diseased joint therefore, it should block the majority of transmission arising from inflammation and/or joint damage driven nociception^6^. We hypothesized that the level of pain reported post-injection might differentiate those patients with predominantly centrally, versus peripherally, mediated pain. In the absence of studies validating this use of lidocaine in IA we carried out an extended analysis of the pain experience using patient-reported questionnaires and quantitative sensory testing to study the integrity of central nervous system pain processing circuits in a group of patients.

PainDETECT, originally established as a validated screening tool to detect possible neuropathic pain components in patients with chronic lower back pain^12^, emerged as a means to predict, with high sensitivity, pain type (based on symptoms) and severity. Relevant for our study, the assessed symptoms are not specific to (although more frequent in) neuropathic pain and share features of centrally mediated pain. In our study, all patients had actively inflamed joints so would be expected to have significant peripheral nociceptive pain. We observed reductions in pain scores post lidocaine injection regardless of whether the individual scored high (>18) or low (≤18) when completing the painDETECT questionnaire, possibly reflecting a similar degree of peripherally mediated pain in all patients. However, those with high painDETECT scores reported greater ongoing NRS pain scores after intra-articular lidocaine injection compared to patients with low painDETECT scores.

Although central pain mechanisms are classically described as a consequence of ongoing nociceptive input^5^ (for example following nerve injury or inflammation) it is increasingly recognised that centrally mediated pain can result independently of peripheral input. Mechanisms that underpin centrally mediated pain are not likely to reverse within the 10-minute timeframe that our post injection pain scores were collected, thus, we propose that people with high ongoing pain scores despite lidocaine administration are experiencing pain that is mechanistically underpinned by pro-nociceptive central processes.

Crucially, painDETECT as a screening tool to detect possible elements of neuropathic pain in a disease state does not include a physical examination and it is noteworthy that, although the painDETECT questionnaire has been used in the literature to indicate central pain processes^8,19,20^, validation against quantitative sensory testing or measures of inferred underlying mechanisms in inflammatory arthritis is limited^21^. To address these limitations, we broadened our analysis in a second ‘validation’ cohort including more measures indicative of maladaptive central nervous system plasticity in chronic pain states: fibromyalgia criteria, in addition to static and dynamic QST methods. Interestingly, individuals 1) stratified into the high painDETECT group 2) meeting fibromyalgia criteria and 3) displaying low non-articular pressure pain thresholds (PPT) i.e. at the trapezius, reported higher pain NRS scores post intra-articular lidocaine. The use of static QST testing provides an inference of functionality in nervous system primary afferent fibres, where PPT measures, for example, may provide an indication of sensory gain (hyperesthesia, hyperalgesia, allodynia) or loss (hypoesthesia, hypoalgesia) of function, pointing to small and/or large diameter nerve fibre dysfunction according to a psychophysical profile. Static measures also may indicate centrally mediated pain when, as investigated in this study, thresholds deviate from normal at non-diseased sites such as the trapezius.

In contrast, differences in post injection pain NRS levels were not statistically significant when categorized by dynamic QST measures. These included temporal summation of pain (TSP) and conditioned pain modulation (CPM), where TSP paradigm outcomes provide a proxy measure of spinal facilitatory processes, and CPM paradigm outcomes provide a proxy measure of functionality in a modulatory process that, in health, acts to inhibit spinal neuronal activity. Recent meta-analyses of studies of pain mechanisms in IA report questionnaire data^2^ and low PPT at extra-articular sites, suggestive of widespread hyperalgesia^22^, suggesting the presence of centrally mediated pain in IA. In contrast, studies on the roles of spinal facilitatory and descending modulatory mechanism in IA were inconclusive^22^. These results are similar to our lack of finding with regards to responses to intra-articular lidocaine when patients are grouped by TSP and CPM responses but align with questionnaire and PPT data. Our results, which demonstrate lowered PPT at the trapezius with no indication of abnormal spinal facilitatory/brain modulatory processing, highlight that considering QST outcomes in isolation is a mistake if a mechanistic understanding of a pain-driving centrally mediated process is sought.

This study has limitations. Using our study design, it is impossible to be certain that lidocaine penetrated all nociceptors in the joint, such as those that are in the subchondral bone. However, studies to date on the effects of intra-articular lidocaine administration in OA knees suggest otherwise. Specifically, results have indicated that lidocaine does act on relevant nociceptors in the joint evidenced by both a significant reduction in pain rating according to the visual analogue scale (VAS)^23^ but also higher-pressure pain thresholds at the knee and surrounding muscles^15^. To investigate this more directly, one would need to adopt an alternative method of abolishing peripheral inputs, such as regional lumbar plexus blockade.

## Conclusion

Overall, our findings support the concept that post intra-articular lidocaine pain scores could be used to identify the contribution of central versus peripheral processes. However, no indication of the mechanism(s) underpinning the peripheral or centrally mediated pain may be gleaned from our study.

## Data Availability

All data produced in the present study are available upon reasonable request to the authors

## Acknowledgements

This study is dedicated to Professor Stephen McMahon, whose ideas led to the conception of this study and who was intrinsic to the design of this study.

## Conflict of interest statement

The authors have no conflicts of interest to declare.

## Funding

ZRL (Doctoral Fellowship, NIHR301674) is funded by the National Institute for Health Research (NIHR) for this research project. The views expressed in this publication are those of the author(s) and not necessarily those of the NIHR, NHS or the UK Department of Health and Social Care. FD and LST acknowledge funding from the Wellcome Trust (collaborative award reference 224257/Z/21/Z).

## Registration

This study was pre-registered on www.clinicaltrials.gov prior to first patient enrolment (Identifier NCT05302232, Unique protocol ID 311106). The manuscript is reported in line with CONSORT reporting guidelines.

## Notes

### Competing Interest Statement

The authors have declared no competing interest.

### Clinical Trial

NCT05302232

### Author Declarations

Yorkshire & The Humber-Sheffield research ethics committee (REC reference 22/YH/0051) granted, Bromley research ethics committee and the Health Research Authority (REC 21/LO/0712) granted.

