## Supplementary material for "A randomised controlled trial of the effect of intra-articular lidocaine on pain scores in inflammatory arthritis"

Supplementary Table 1: Amount of steroid and lidocaine/0.9% saline administered to each joint during study visits

| **Joint** | **Steroid (depomedrone 40mg/ml) + 1% lidocaine** | **Steroid (depomedrone 40mg/ml) + 0.9% saline** |
| --- | --- | --- |
| Meta-carpal phalangeal joints | 20mg depomedrone + 0.5ml lidocaine | 10mg depomedrone + 0.5ml saline |
| Wrist, Elbow, Ankle | 40mg depomedrone +1 ml lidocaine | 40mg depomedrone + 1ml saline |
| Knee | 80mg depomedrone + 3mls lidocaine | 80mg depomedrone + 3mls saline |

Standardised amounts of steroid, lidocaine and saline were administered to each joint in order to minimise variation in joint capsule expansion and any effect this may have on pain score.

Supplementary Table 2: NRS pain scores at 5 minutes post intra-articular lidocaine injection

|  | Not centrally mediated | Centrally mediated |
| --- | --- | --- |
| painDETECT (RCT) | 2.8 (2.1) | 5.0 (2.9) |
| painDETECT | 1.9 (2.0) | 4.3 (2.5) |
| Fibromyalgia criteria | 1.7 (1.6) | 4.2 (2.6) |
| PPT at trapezius | 1.8 (1.8) | 3.7 (2.6) |
| Facilitated TSP | 2.1 (2.0) | 2.9 (1.1) |
| Non responder CPM PTT | 2.2 (1.6) | 2.8 (2.6) |

NRS pain scores at 5 minutes post intra-articular lidocaine injection, in those with “centrally mediated” versus “not centrally mediated” according to painDETECT, fibromyalgia criteria, temporal summation of pain facilitation (TSP) or responder status to conditioned pain modulation pressure tolerance threshold (CPM PTT). Shown as mean (SD). Pain NRS score at 5 minutes post intra-articular lidocaine was consistently above 4/10 in the painDETECT high group and fibromyalgia positive group. Abbreviations: RCT (Randomised controlled trial), PPT (Pressure pain threshold), TSP (Temporal summation of pain), CPM PTT (Conditioned pain modulation pressure tolerance threshold). Means shown with SD

Supplementary Table 3: Results of the linear mixed effect model at each of the post-injection time points.

|  |  | **PainDETECT high/low RCT group (n=26)** | **PainDETECT high/low (n=40)** | **FM +ve/-ve (n=40)** | **Trapezius PPT (n=40)** | **Facilitated TSP +ve/-ve (n=32)** | **CPM Responder/Non-responder (n=40)** |
| --- | --- | --- | --- | --- | --- | --- | --- |
| Unadjusted for pre- injection NRS score |  |  |  |  |  |  |  |
|  | At 3 mins | **2.2(0.7-3.7) p=0.004** | **2.9(1.2-4.5) p=0.001** | **2.7(1-4.3) p=0.001** | **1.8 (0.2-3.4)**  **P=0.027** | -2 (-4.8-0.8)  P=0.154 | -0.5(-2.0-1.2) p=0.541 |
|  | At 5 mins | **2.1(0.7-3.5) p=0.003** | **2.4(0.9-3.9) p=0.002** | **2.5 (1.1-3.9) p=0.001** | **1.9 (0.5-3.2)**  **P=0.006** | -1.5 (-3.9-0.9)  P=0.223 | -0.6(-2-0.7) p=0.361 |
|  | At 10 mins | **2.1(0.8-3.4) p=0.002** | **2.1(0.6-3.7) p=0.006** | **2.1 (0.7-3.5) p=0.003** | **2.0 (0.8-3.3)**  **P=0.001** | -1.4 (-3.6-0.9)  P=0.234 | -0.9(-2.2-0.3) p=0.151 |
| Adjusted for pre-injection NRS score |  |  |  |  |  |  |  |
|  | At 3 mins | 0.5(-0.9-2.0) p=0.46 | **2.3(1-3.5) p=0.000** | **1.9 (0.4-3.4) p=0.015** | 1.1 (-0.3-2.5)  P=0.1 | -1.9 (-4.2-0.3)  P=0.093 | -0.0(-1.1-1.1) p=0.988 |
|  | At 5 mins | 0.4(-0.9-1.8) p=0.53 | **1.8(0.7-2.9) p=0.002** | **1.7(0.4-3) p=0.013** | 1.2 (-0.1-2.4)  P=0.06 | -1.4 (-3.4-0.5)  P=0.144 | -0.1(-1.2-0.9) p=0.792 |
|  | At 10 mins | 0.4(-0.8-1.7) p=0.51 | **1.6(0.4-2.7) p=0.002** | **1.3(0.03-2.6) p=0.045** | **1.3 (0.2-2.5)**  **P=0.02** | -1.3 (-3.1-0.5)  P=0.146 | -0.5(-1.7-0.8) p=0.457 |

Results of the linear mixed effects model showing mean difference (95% confidence intervals) in NRS pain scores at each post intervention time point (3, 5 and 10 minutes), for patients grouped according to measures of centrally mediated pain. Those shown in bold are significant at the 5% level. FM= fulfillment of fibromyalgia criteria, PPT= pressure pain threshold, TSP= temporal summation of pain, CPM= conditioned pain modulation

Supplementary Table 4: Demographics of PUMIA patients, the second study population.

| **Variable** | **Low painDETECT (n= 27)** | **High painDETECT (n= 13)** | **Total (n=40)** | **Significance** |
| --- | --- | --- | --- | --- |
| Age, years (Mean, SD) | 50 (15 ) | 58 (14) | 52.7 (15) | P=0.112 |
| Gender, F (N, %) | 18 (67%) | 11 (85%) | 29 (73%) |  |
| Diagnosis, (N, %)  EIA  Rheumatoid Arthritis  PsA  Peripheral SpA | 4 (15%)  20 (74%)  2 (7%)  1 (4%) | 0 (0%)  13 (100%)  0 (0%)  0 (0%) | 4 (10%)  33 (83%)  2 (5%)  1 (3%) |  |
| Symptom duration, years (Mean, SD) | 6.8 (10) | 9.8 (10) | 7.8 (10) | P=0.394 |
| DAS28 (Mean, SD) | 4.3 (1.2) | 5.3 (2.4) | 4.6 (1.2) | P<0.05 |
| MSK HQ score (Mean, SD) | 29 (12) | 17 (8) | 24.7 (12) | P<0.05 |
| PHQ9 (Mean, SD) | 7.2 (6) | 14.6 (6) | 9.6 (6.9) | P<0.05 |
| GAD7 (Mean, SD) | 5.7 (6) | 12.5 (7) | 7.9 (6.9) | P<0.05 |
| PHQ15 (Mean, SD) | 6.8 (4) | 11.8 (4) | 8.5 (4.4) | P<0.05 |
| Pre injection pain NRS score (Mean, SD) | 5.9 (2.4) | 7.1 (2.6) | 6.3 (2.5) | P=0.193 |
| WPI (Mean, SD) | 5 (2.5) | 9.7 (4.3) | 6.5 (3.4) | P<0.05 |
| SSS (Mean, SD) | 4.4 (1.2) | 8.5 (2.5) | 5.8 (3.0) | P<0.05 |
| Joint  Knee  Wrist  Elbow  Ankle  MCP | 6 (22%)  14 (52%)  3 (11%)  2 (7%)  2 (7%) | 3 (23%)  9 (69%)  0 (0%)  0 (0%)  1 (8%) | 9 (23%)  23 (58%)  3 (8%)  2 (5%)  3 (8%) |  |
| Ultrasound at target joint  GS +ve (1-3)  PD +ve (1-3) | 25 (93%)  25 (93%) | 10 (77%)  8 (62%) | 35 (88%)  33 (83%) |  |
| painDETECT score (Mean, SD) | 11 (4.5) | 25 (3.5) | 15 (7.8) | P<0.05 |
| PPT at trapezius Kg/cm^2^ (Median) | 2.1 | 2.2 | 2.1 |  |

Demographics of second study population, PUMIA participants, shown as demographics of those with low painDETECT and those with high painDETECT and then total population. Those with high painDETECT had significantly greater disease activity (DAS 28), lower quality of life (shown by lower MSK HQ score), higher levels of depression (PHQ9), higher anxiety (GAD7), higher somatisation (PHQ15) and higher scores on fibromyalgia criteria (WPI and SSS). Abbreviations: DAS28 (Disease activity score 28), MSK HQ (Musculoskeletal health questionnaire), PHQ9 (Patient health questionnaire 9), GAD 7 (Generalised anxiety disorder assessment 7), PHQ15 (Patient health questionnaire 15), WPI (Widespread pain index), SSS (Symptom severity scale), GS (Grey-scale), PD (Power doppler), PPT (Pressure pain threshold).

Supplementary Figure 1: Drop in NRS pain score pre and post placebo control, grouped by painDETECT low/high in RCT group.

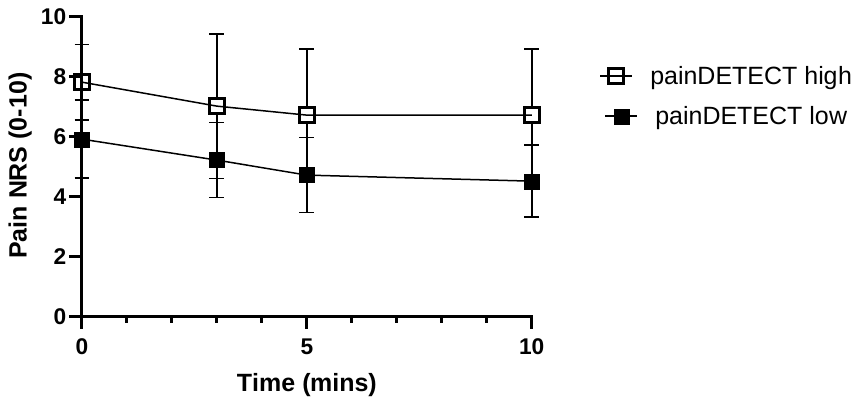
Supplementary Figure 1: Pain NRS response at 3,5 and 10 minutes post to intra-articular control injection in RCT cohort, grouped according to painDETECT high/low. (mean ± 95% CI).
